# Trends and variation in andexanet alfa for the reversal of direct oral anticoagulants in NHS Trusts in England

**DOI:** 10.1101/2025.01.21.25320733

**Authors:** Louis Fisher, Richard Buka, Rosalind Byrne, Stephen Black, Helen J Curtis, Christopher Wood, Andrew Brown, Sebastian Bacon, Richard Croker, Ben Goldacre, Brian MacKenna, Victoria Speed

## Abstract

Andexanet alfa was recommended by the National Institute for Health and Clinical Excellence (NICE) as an option in the management of major gastrointestinal bleeding in patients taking apixaban or rivaroxaban in May 2021. To assess the uptake of andexanet alfa, we analysed pharmacy stock control data from NHS Trusts in England using the openly available Secondary Care Medicines Dataset.

Between May 2021 and July 2024, 14,092 vials of andexanet alfa were issued in NHS Trusts in England. There was wide variation in uptake across NHS Trusts, which likely reflects variation in local protocols.

This study is the first analysis of adoption of a new treatment using the OpenPrescribing Hospitals platform, which is expected to provide a generalisable framework for similar analyses in the future.

## Introduction

The National Institute for Health and Clinical Excellence (NICE) recommends andexanet alfa (AA) as an option in the management of major gastrointestinal (GI) bleeding in patients taking apixaban or rivaroxaban (NICE TA697, May 2021).(1) The recommendation was based on findings from the single-arm ANNEXA-4 trial where 90 patients with major GI bleeding received AA and 85% were found to have good or excellent haemostatic efficacy.(1,2) A subsequent trial, ANNEXA-I, randomised patients with DOAC-associated intracerebral haemorrhage to AA or usual care, and found a reduction in haematoma expansion but no improvement in clinical outcomes including death or disability(3) however the trial was not powered for these outcomes. Importantly, both trials show that AA increases the risk of thrombosis, predominantly arterial.

Andexanet alfa is also expensive.(4) The National Health Service (NHS) list price is approximately £15,000 per patient. Whilst the NHS receives a discount for AA, the amount is commercially sensitive and confidential.(1) In this context, a recent UK-wide survey of local hospital protocols for the use of AA suggested that practice across the UK is highly variable.(5)

Although primary care prescribing data has been extensively used for variation in care analysis, until recently, there was no open access to hospital medicines usage data. As this dataset has become newly available we set out to report the volume of AA issued in secondary care trusts in England, and to report regional trends and variation.

## Methods

Secondary Care Medicines Data (SCMD), collated by Rx-info, was accessed using the NHS Business Services Authority (NHSBSA).(6) This data contains issued pharmacy stock control data from all NHS Acute, Teaching, Specialist, Mental Health and Community Trusts in England. Individual Trusts are represented using Organisational Data Service (ODS) codes, which were mapped to NHS regions using files provided by the ODS. Using open data, we identified Trusts with 24-hour consultant-led emergency care activity reported in the last six months.(7) Andexanet alfa was identified in the SCMD using the Systematized Nomenclature of Medicine - Clinical Term (SNOMED CT) ‘37454211000001101’.(8) We counted the number of vials of AA issued at a regional and Trust level. We calculated a six-month rolling average of the monthly quantity of issued vials between May 2021 and July 2024 broken down by NHS region and Trust-level deciles for the monthly quantity of issued vials. Both were plotted as time series charts. For context, the recommended dose of AA is five vials for the *low dose* regimen or nine vials for the *high dose* regimen.(9)

All data are openly available.(6) No ethical approval was sought for this analysis. Code for data management and analysis, are openly available for inspection and re-use at <u>github.com/ebmdatalab/op-hospitals-andexanet</u>.

## Results

14,092 vials of AA were issued between May 2021 and July 2024. 115/119 (96.6%) eligible Trusts issued AA in the study period (**Table S1**).

Figure 1 shows the 6-month rolling average in each of the seven NHS regions in England. There is regional variation in the total issued and speed of uptake. From May 2022, most AA was issued in the Midlands. Assuming all vials had been administered, between 1,579 (14,205/9 vials, high dose regimen) and 2,841 (14,205/5 vials, low dose regimen) patients have been treated to date in England.

**Figure 1.**
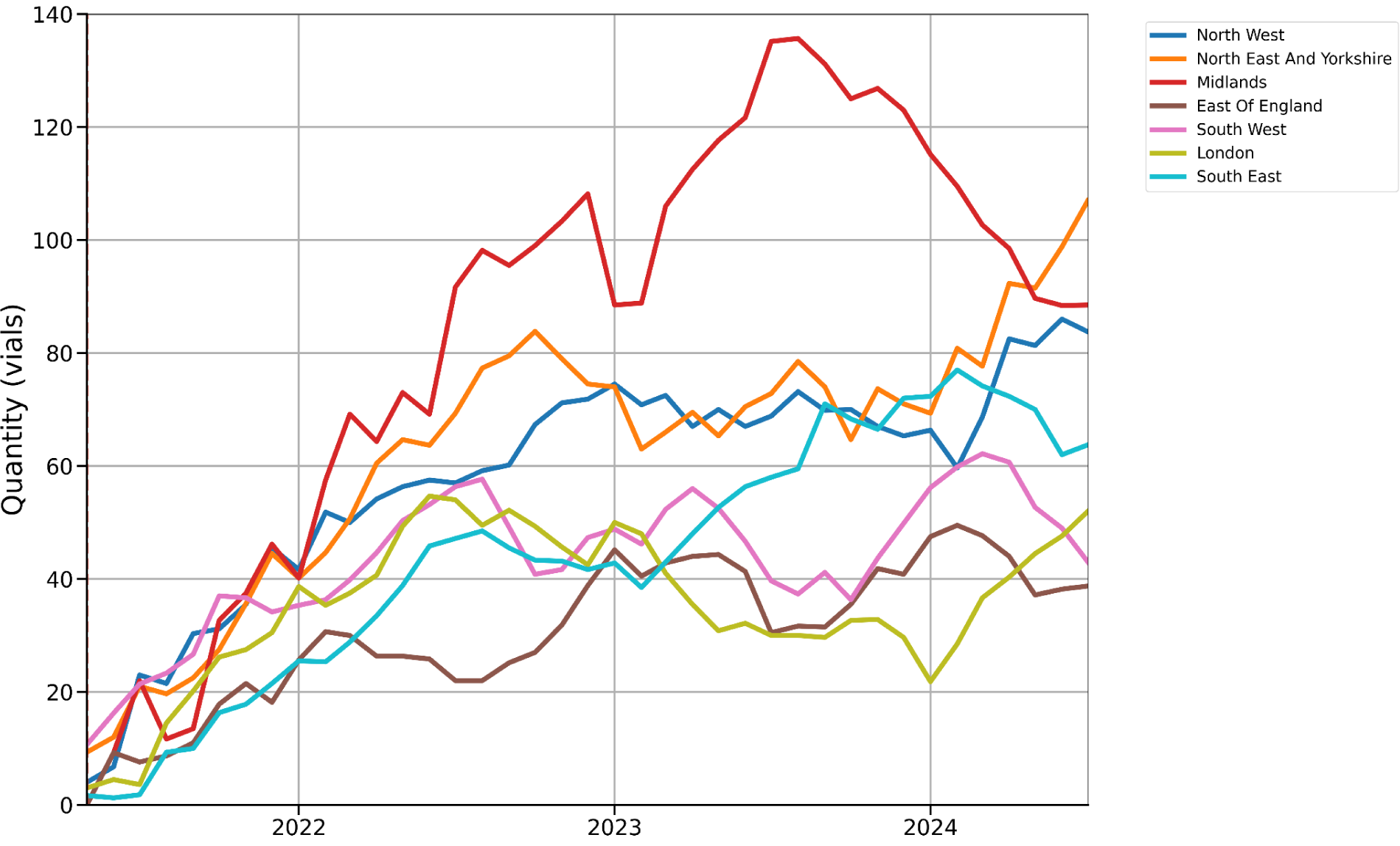
The monthly number of vials of AA issued in secondary care between May 2021 and July 2024 by NHS region (6-month rolling average).

There was variation in uptake amongst NHS Trusts (Figure 2, note for interpretation of decile ranges that the Trust median is zero until mid 2024). Total usage since May 2021 for the 25 Trusts with most usage is reported in **Figure S1.**

**Figure 2.**
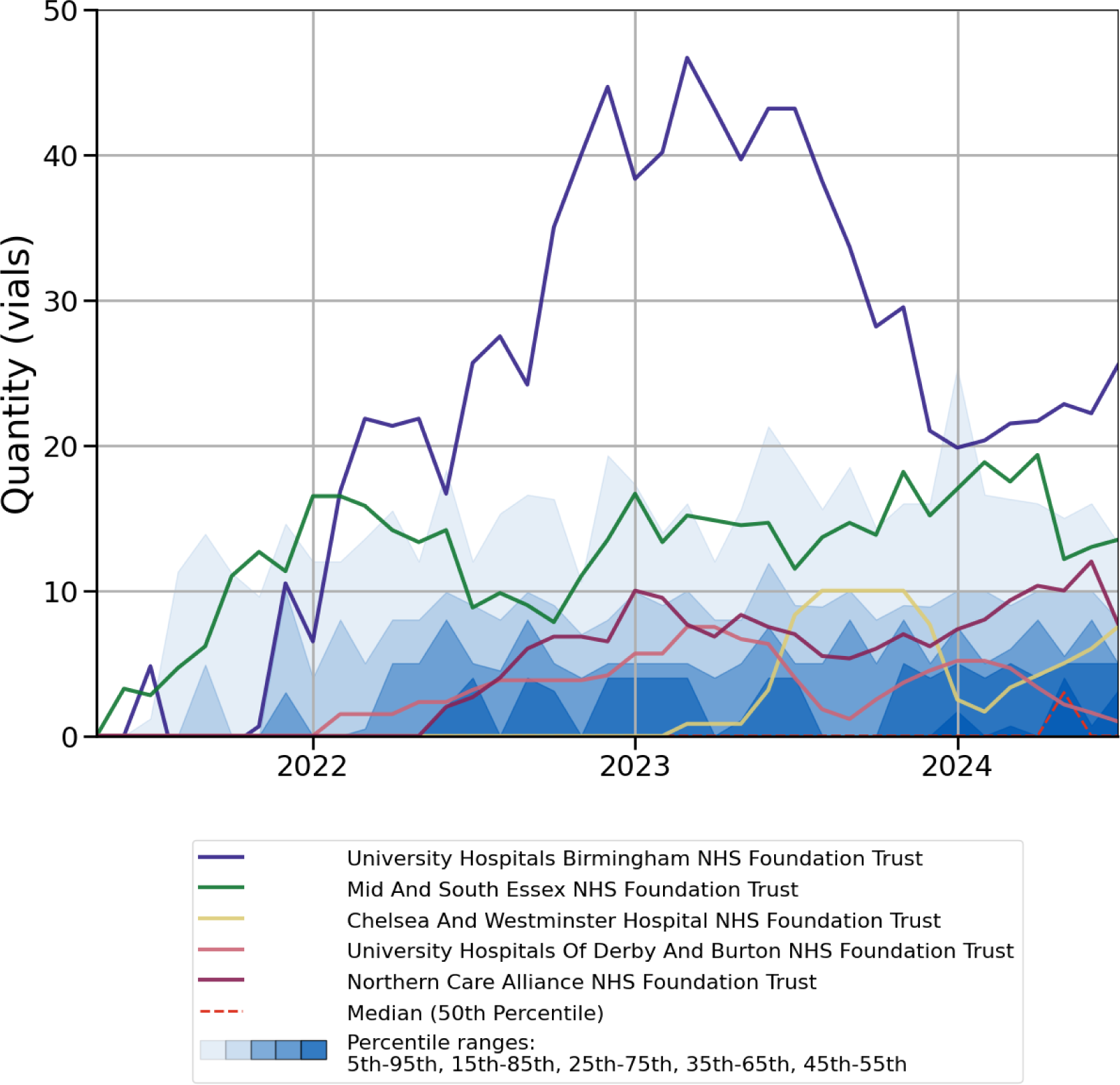
The monthly number of vials of AA issued in secondary care in five large NHS Trusts between May 2021 and July 2024. The shaded deciles represent variation in monthly counts of AA vials issued across all included Trusts. The individual Trusts are reported as a 6 month rolling average of the monthly counts of AA vials issued.

## Discussion

This is the first study to describe the marked variation in adoption of AA in England. Our data demonstrate widespread uptake of AA since the publication of the NICE TA in May 2021 and are consistent with AA being present on the Royal College of Emergency Physicians Antidote list(10); but also suggest widespread low or non-usage in many Trusts. It is the first publication from the new <u>OpenPrescribing Hospitals</u> platform, a new openly accessible project to enable anyone to analyse medicines issued in NHS Trusts England using open data; it builds on the successful delivery of <u>openprescribing.net</u>, an analytic platform for prescribing data in primary care with 20,000 users per month.(11)

The openly available SCMD is a rich data source. However, there are limitations in the use of stock control data where patient level medicines usage is not available. For this reason, we cannot determine whether the AA has been used for indications outside NICE recommendations, for example intracranial haemorrhage. Where a Trust has not issued AA, we cannot be sure whether this because it has not been issued yet (eg. not been prescribed for a patient) or whether the Trust does not hold stock. Similarly we were unable to differentiate when AA was issued but not administered (eg. expired). Further, we necessarily report our findings without denominators (as these cannot be created for secondary care Trusts); however the count data alone provides a helpful insight into uptake. Finally, actual cost is not reported in the SCMD (only indicative cost, which does not reflect the discounts applied by the manufacturer for the NHS).

The study highlights considerable variation in the uptake of a new, expensive agent. This wide variation is likely due to variation in local protocols which are influenced by individual clinicians’ interpretation of the evidence(5). This raises important questions for policy makers and those responsible for local clinical guidance implementation. For clarity on the risk and benefit of AA for GI haemorrhage a randomised controlled trial is needed: until a trial is commissioned, the natural variation identified here may provide an opportunity to estimate effectiveness, using treating hospital as an instrumental variable.

This study provides the first window into the adoption of a new high cost treatment by NHS hospitals, from a platform that is expected to produce a generalised service describing varied adoption of many such treatments in the future. Further research is required to understand the reasons why AA is used heavily in some centres and not at all in others.

## Acknowledgements

### Funding

This work was supported by The NIHR Biomedical Research Centre, Oxford, A Health Foundation grant (Award Reference Number 7599); A National Institute for Health Research (NIHR) School of Primary Care Research (SPCR) grant (Award Reference Number 327); the National Institute for Health Research (NIHR) under its Research for Patient Benefit (RfPB) Programme (Grant Reference Number PB-PG-0418-20036) and by the National Institute for Health Research Applied Research Collaboration Oxford and Thames Valley. The views expressed in this publication are those of the author(s) and not necessarily those of the NIHR, NHS England or the Department of Health and Social Care. Funders had no role in the study design, collection, analysis, and interpretation of data; in the writing of the report; and in the decision to submit the article for publication. The development of the OpenPrescribing Hospitals software has been funded by the NHS Primary Care and Medicines Analytics Unit.

### Ethical approval

This study uses exclusively open, publicly available data, therefore no ethical approval was required.

### Guarantor

VS is guarantor.

### Contributorship

**Conceptualization**: Louis Fisher and Victoria Speed.

**Data curation**: Louis Fisher and Stephen Black.

**Formal analysis**: Louis Fisher and Victoria Speed.

**Funding acquisition**: Ben Goldacre and Brian MacKenna.

**Investigation**: Louis Fisher, Stephen Black, and Victoria Speed.

**Methodology**: Louis Fisher, Stephen Black, Richard Croker, Ben Goldacre, and Brian MacKenna.

**Project administration**: Victoria Speed.

**Resources**: Sebastian Bacon.

**Software**: Louis Fisher and Sebastian Bacon. Supervision: Ben Goldacre.

**Validation**: Louis Fisher, Christopher Wood, Andrew Brown, Richard Croker, and Victoria Speed.

**Visualization**: Louis Fisher, Ben Goldacre, and Victoria Speed.

**Writing - original draft**: Louis Fisher, Richard Buka, Rosalind Byrne, and Victoria Speed. **Writing - review & editing**: Louis Fisher, Richard Buka, Rosalind Byrne, Stephen Black, Helen J. Curtis, Christopher Wood, Andrew Brown, Sebastian Bacon, Richard Croker, Ben Goldacre, Brian MacKenna, and Victoria Speed.

## Supporting information

STROBE checklist

## Data Availability

The data, code and results for this study are openly available.

https://opendata.nhsbsa.net/dataset/secondary-care-medicines-data-indicative-price

http://github.com/ebmdatalab/op-hospitals-andexanet

## References

1. Overview | Andexanet alfa for reversing anticoagulation from apixaban or rivaroxaban | Guidance | NICE. [cited 2024 Aug 14]; Available from: https://www.nice.org.uk/guidance/ta697

2. Connolly Stuart J., Crowther Mark, Eikelboom John W., Gibson C. Michael, Curnutte John T., Lawrence John H., et al. Full Study Report of Andexanet Alfa for Bleeding Associated with Factor Xa Inhibitors. N Engl J Med. 2019 Apr 4;380(14):1326–35.

3. Connolly Stuart J., Sharma Mukul, Cohen Alexander T., Demchuk Andrew M., Członkowska Anna, Lindgren Arne G., et al. Andexanet for Factor Xa Inhibitor–Associated Acute Intracerebral Hemorrhage. N Engl J Med. 2024 May 15;390(19):1745–55.

4. British National Formulary, andexanet alfa [Internet]. [cited 2024 Sep 9]. Available from: https://bnf.nice.org.uk/drugs/andexanet-alfa/medicinal-forms/

5. Glancy P, Sutton DJ, Gomez K, Nicolson PLR, Buka RJ. How will UK hospitals use andexanet alfa? A review of local protocols. EJHaem. 2023 Feb;4(1):298–300.

6. NHS Business Services Authority. Secondary Care Medicines Data (SCMD) with indicative price. Open Data Portal (NHS Business Services Authority). Available from: https://opendata.nhsbsa.net/dataset/secondary-care-medicines-data-indicative-price.

7. England NHS. A&E Attendances and Emergency Admissions [Internet]. [cited 2024 Oct 2]. Available from: https://www.england.nhs.uk/statistics/statistical-work-areas/ae-waiting-times-and-activity/

8. OpenPrescribing dm+d browser [Internet]. [cited 2024 Oct 7]. Andexanet alfa 200mg powder for solution for infusion vials. Available from: https://openprescribing.net/dmd/vmp/37454211000001101/

9. Ondexxya 200 mg powder for solution for infusion - Summary of Product Characteristics (SmPC) - (emc) [Internet]. [cited 2024 Oct 7]. Available from: https://www.medicines.org.uk/emc/product/10933/smpc#:~:text=Andexanet%20alfa%20is%20a%20specific%20reversal

10. Draper S. RCEM. 2021 [cited 2024 Sep 9]. RCEM NPIS Antidote Guideline Update 2021. Available from: https://rcem.ac.uk/rcem-npis-antidote-guideline-update-2021/

11. Home [Internet]. [cited 2024 Nov 20]. Available from: https://openprescribing.net/

