## Supplementary material for "Trends and variation in andexanet alfa for the reversal of direct oral anticoagulants in NHS Trusts in England"

### Figure S1. Top 25 NHS Trusts by number of vials of andexanet alfa used between May 2021 and July 2024

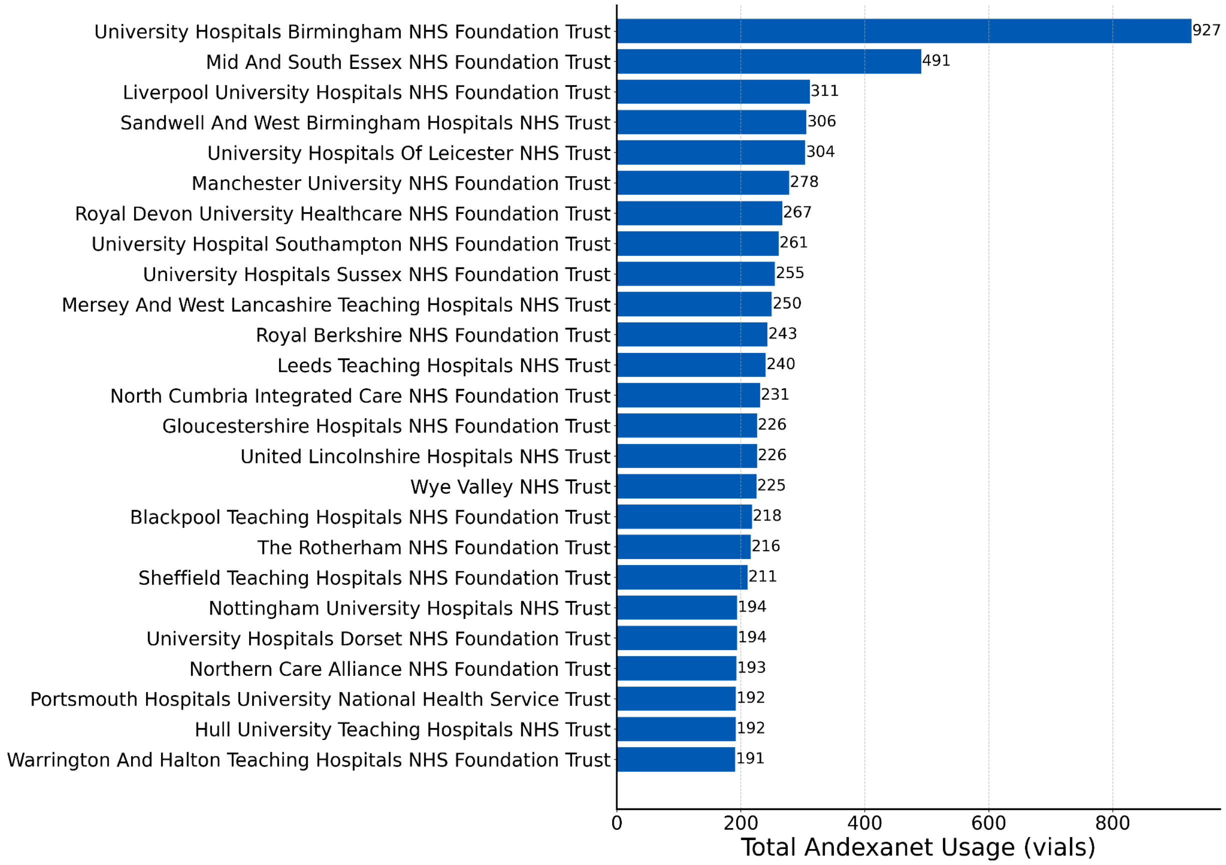

**Supplementary Table 1.** NHS Trusts in the SCMD, with an indication of whether they had any emergency department activity in the last 6 months (and were therefore included in the analysis) and an indication of whether they issued any andexanet alfa between May 2021 and July 2024.

| ODS code | Trust name | Emergency department activity in the last 6 months* | AA issued |
|---|---|---|---|
| RCF | Airedale NHS Foundation Trust | Yes | Yes |
| RTK | Ashford And St Peter'S Hospitals NHS Foundation Trust | Yes | Yes |
| RF4 | Barking, Havering And Redbridge University Hospitals NHS Trust | Yes | Yes |
| RFF | Barnsley Hospital NHS Foundation Trust | Yes | Yes |
| R1H | Barts Health NHS Trust | Yes | Yes |
| RC9 | Bedfordshire Hospitals NHS Foundation Trust | Yes | Yes |
| RXL | Blackpool Teaching Hospitals NHS Foundation Trust | Yes | Yes |
| RMC | Bolton NHS Foundation Trust | Yes | Yes |
| RAE | Bradford Teaching Hospitals NHS Foundation Trust | Yes | Yes |
| RXQ | Buckinghamshire Healthcare NHS Trust | Yes | Yes |
| RWY | Calderdale And Huddersfield NHS Foundation Trust | Yes | Yes |
| RGT | Cambridge University Hospitals NHS Foundation Trust | Yes | Yes |
| RQM | Chelsea And Westminster Hospital NHS Foundation Trust | Yes | Yes |
| RFS | Chesterfield Royal Hospital NHS Foundation Trust | Yes | Yes |
| RJR | Countess Of Chester Hospital NHS Foundation Trust | Yes | Yes |
| RXP | County Durham And Darlington NHS Foundation Trust | Yes | Yes |
| RJ6 | Croydon Health Services NHS Trust | Yes | Yes |
| RN7 | Dartford And Gravesham NHS Trust | Yes | Yes |
| RP5 | Doncaster And Bassetlaw Teaching Hospitals NHS Foundation Trust | Yes | Yes |

| | | | |
|---|---|---|---|
| RBD | Dorset County Hospital NHS Foundation Trust | Yes | Yes |
| RWH | East And North Hertfordshire NHS Trust | Yes | Yes |
| RJN | East Cheshire NHS Trust | Yes | Yes |
| RVV | East Kent Hospitals University NHS Foundation Trust | Yes | Yes |
| RXR | East Lancashire Hospitals NHS Trust | Yes | Yes |
| RDE | East Suffolk And North Essex NHS Foundation Trust | Yes | Yes |
| RXC | East Sussex Healthcare NHS Trust | Yes | Yes |
| RDU | Frimley Health NHS Foundation Trust | Yes | Yes |
| RR7 | Gateshead Health NHS Foundation Trust | Yes | Yes |
| RLT | George Eliot Hospital NHS Trust | Yes | Yes |
| RTE | Gloucestershire Hospitals NHS Foundation Trust | Yes | Yes |
| RN3 | Great Western Hospitals NHS Foundation Trust | Yes | Yes |
| RJ1 | Guy's And St Thomas' NHS Foundation Trust | Yes | Yes |
| RN5 | Hampshire Hospitals NHS Foundation Trust | Yes | Yes |
| RCD | Harrogate And District NHS Foundation Trust | Yes | Yes |
| RWA | Hull University Teaching Hospitals NHS Trust | Yes | Yes |
| RYJ | Imperial College Healthcare NHS Trust | Yes | Yes |
| R1F | Isle Of Wight NHS Trust | Yes | Yes |
| RGP | James Paget University Hospitals NHS Foundation Trust | Yes | Yes |
| RNQ | Kettering General Hospital NHS Foundation Trust | Yes | Yes |
| RJZ | King's College Hospital NHS Foundation Trust | Yes | Yes |
| RAX | Kingston Hospital NHS Foundation Trust | Yes | Yes |
| RXN | Lancashire Teaching Hospitals NHS Foundation Trust | Yes | Yes |
| RR8 | Leeds Teaching Hospitals NHS Trust | Yes | Yes |
| RJ2 | Lewisham And Greenwich NHS Trust | Yes | Yes |

| | | | |
|---|---|---|---|
| REM | Liverpool University Hospitals NHS Foundation Trust | Yes | Yes |
| R1K | London North West University Healthcare NHS Trust | Yes | Yes |
| RWF | Maidstone And Tunbridge Wells NHS Trust | Yes | Yes |
| R0A | Manchester University NHS Foundation Trust | Yes | Yes |
| RPA | Medway NHS Foundation Trust | Yes | Yes |
| RBN | Mersey And West Lancashire Teaching Hospitals NHS Trust | Yes | Yes |
| RAJ | Mid And South Essex NHS Foundation Trust | Yes | Yes |
| RBT | Mid Cheshire Hospitals NHS Foundation Trust | Yes | Yes |
| RXF | Mid Yorkshire Teaching NHS Trust | Yes | Yes |
| RD8 | Milton Keynes University Hospital NHS Foundation Trust | Yes | Yes |
| RM1 | Norfolk And Norwich University Hospitals NHS Foundation Trust | Yes | Yes |
| RVJ | North Bristol NHS Trust | Yes | Yes |
| RNN | North Cumbria Integrated Care NHS Foundation Trust | Yes | Yes |
| RAP | North Middlesex University Hospital NHS Trust | Yes | Yes |
| RVW | North Tees And Hartlepool NHS Foundation Trust | Yes | Yes |
| RNS | Northampton General Hospital NHS Trust | Yes | Yes |
| RM3 | Northern Care Alliance NHS Foundation Trust | Yes | Yes |
| RJL | Northern Lincolnshire And Goole NHS Foundation Trust | Yes | Yes |
| RTF | Northumbria Healthcare NHS Foundation Trust | Yes | Yes |
| RX1 | Nottingham University Hospitals NHS Trust | Yes | Yes |
| RHU | Portsmouth Hospitals University National Health Service Trust | Yes | Yes |
| RHW | Royal Berkshire NHS Foundation Trust | Yes | Yes |
| REF | Royal Cornwall Hospitals NHS Trust | Yes | Yes |

| | | | |
|---|---|---|---|
| RH8 | Royal Devon University Healthcare NHS Foundation Trust | Yes | Yes |
| RAL | Royal Free London NHS Foundation Trust | Yes | Yes |
| RA2 | Royal Surrey County Hospital NHS Foundation Trust | Yes | Yes |
| RD1 | Royal United Hospitals Bath NHS Foundation Trust | Yes | Yes |
| RNZ | Salisbury NHS Foundation Trust | Yes | Yes |
| RXK | Sandwell And West Birmingham Hospitals NHS Trust | Yes | Yes |
| RHQ | Sheffield Teaching Hospitals NHS Foundation Trust | Yes | Yes |
| RK5 | Sherwood Forest Hospitals NHS Foundation Trust | Yes | Yes |
| RH5 | Somerset NHS Foundation Trust | Yes | Yes |
| RTR | South Tees Hospitals NHS Foundation Trust | Yes | Yes |
| R0B | South Tyneside And Sunderland NHS Foundation Trust | Yes | Yes |
| RJC | South Warwickshire University NHS Foundation Trust | Yes | Yes |
| RJ7 | St George'S University Hospitals NHS Foundation Trust | Yes | Yes |
| RWJ | Stockport NHS Foundation Trust | Yes | Yes |
| RTP | Surrey And Sussex Healthcare NHS Trust | Yes | Yes |
| RMP | Tameside And Glossop Integrated Care NHS Foundation Trust | Yes | Yes |
| RNA | The Dudley Group NHS Foundation Trust | Yes | Yes |
| RAS | The Hillingdon Hospitals NHS Foundation Trust | Yes | Yes |
| RTD | The Newcastle Upon Tyne Hospitals NHS Foundation Trust | Yes | Yes |
| RQW | The Princess Alexandra Hospital NHS Trust | Yes | Yes |
| RCX | The Queen Elizabeth Hospital, King'S Lynn, NHS Foundation Trust | Yes | Yes |
| RFR | The Rotherham NHS Foundation Trust | Yes | Yes |
| RL4 | The Royal Wolverhampton NHS Trust | Yes | Yes |
| RXW | The Shrewsbury And Telford Hospital NHS Trust | Yes | Yes |

| | | | |
|---|---|---|---|
| RA9 | Torbay And South Devon NHS Foundation Trust | Yes | Yes |
| RWD | United Lincolnshire Hospitals NHS Trust | Yes | Yes |
| RRV | University College London Hospitals NHS Foundation Trust | Yes | Yes |
| RHM | University Hospital Southampton NHS Foundation Trust | Yes | Yes |
| RRK | University Hospitals Birmingham NHS Foundation Trust | Yes | Yes |
| RA7 | University Hospitals Bristol And Weston NHS Foundation Trust | Yes | Yes |
| RKB | University Hospitals Coventry And Warwickshire NHS Trust | Yes | Yes |
| R0D | University Hospitals Dorset NHS Foundation Trust | Yes | Yes |
| RTG | University Hospitals Of Derby And Burton NHS Foundation Trust | Yes | Yes |
| RWE | University Hospitals Of Leicester NHS Trust | Yes | Yes |
| RTX | University Hospitals Of Morecambe Bay NHS Foundation Trust | Yes | Yes |
| RJE | University Hospitals Of North Midlands NHS Trust | Yes | Yes |
| RK9 | University Hospitals Plymouth NHS Trust | Yes | Yes |
| RYR | University Hospitals Sussex NHS Foundation Trust | Yes | Yes |
| RBK | Walsall Healthcare NHS Trust | Yes | Yes |
| RWW | Warrington And Halton Teaching Hospitals NHS Foundation Trust | Yes | Yes |
| RWG | West Hertfordshire Teaching Hospitals NHS Trust | Yes | Yes |
| RGR | West Suffolk NHS Foundation Trust | Yes | Yes |
| RKE | Whittington Health NHS Trust | Yes | Yes |
| RBL | Wirral University Teaching Hospital NHS Foundation Trust | Yes | Yes |
| RWP | Worcestershire Acute Hospitals NHS Trust | Yes | Yes |
| RRF | Wrightington, Wigan And Leigh NHS Foundation Trust | Yes | Yes |
| RLQ | Wye Valley NHS Trust | Yes | Yes |

| | | | |
|---|---|---|---|
| RCB | York And Scarborough Teaching Hospitals NHS Foundation Trust | Yes | Yes |
| RBS | Alder Hey Children's NHS Foundation Trust | Paediatric (not included) | No |
| RQ3 | Birmingham Women's And Children'S NHS Foundation Trust | Paediatric and womens (not included) | No |
| RVR | Epsom And St Helier University Hospitals NHS Trust | Yes | No |
| RQX | Homerton Healthcare NHS Foundation Trust | Yes | No |
| RGN | North West Anglia NHS Foundation Trust | Yes | No |
| RTH | Oxford University Hospitals NHS Foundation Trust | Yes | No |
| RCU | Sheffield Children's NHS Foundation Trust | Paediatric (not included) | No |
| RBV | The Christie NHS Foundation Trust | No | Yes |
| RVN | Avon And Wiltshire Mental Health Partnership NHS Trust | No | No |
| RRP | Barnet, Enfield And Haringey Mental Health NHS Trust | No | No |
| RV7 | Bedfordshire And Luton Mental Health And Social Care Partnership NHS Trust | No | No |
| RWX | Berkshire Healthcare NHS Foundation Trust | No | No |
| RXT | Birmingham And Solihull Mental Health NHS Foundation Trust | No | No |
| RYW | Birmingham Community Healthcare NHS Foundation Trust | No | No |
| TAJ | Black Country Healthcare NHS Foundation Trust | No | No |
| TAD | Bradford District Care NHS Foundation Trust | No | No |
| RY2 | Bridgewater Community Healthcare NHS Foundation Trust | No | No |
| RXH | Brighton And Sussex University Hospitals NHS Trust | No | No |
| RT1 | Cambridgeshire And Peterborough NHS Foundation Trust | No | No |
| RYV | Cambridgeshire Community Services NHS Trust | No | No |

| | | | |
|---|---|---|---|
| TAF | Camden And Islington NHS Foundation Trust | No | No |
| RV3 | Central And North West London NHS Foundation Trust | No | No |
| RYX | Central London Community Healthcare NHS Trust | No | No |
| RXA | Cheshire And Wirral Partnership NHS Foundation Trust | No | No |
| RJ8 | Cornwall Partnership NHS Foundation Trust | No | No |
| RYG | Coventry And Warwickshire Partnership NHS Trust | No | No |
| RX4 | Cumbria, Northumberland, Tyne And Wear NHS Foundation Trust | No | No |
| RY8 | Derbyshire Community Health Services NHS Foundation Trust | No | No |
| RXM | Derbyshire Healthcare NHS Foundation Trust | No | No |
| RWV | Devon Partnership NHS Trust | No | No |
| RDY | Dorset Healthcare University NHS Foundation Trust | No | No |
| RYK | Dudley Integrated Health And Care NHS Trust | No | No |
| RWK | East London NHS Foundation Trust | No | No |
| RX9 | East Midlands Ambulance Service NHS Trust | No | No |
| RYC | East Of England Ambulance Service NHS Trust | No | No |
| R1L | Essex Partnership University NHS Foundation Trust | No | No |
| RTQ | Gloucestershire Health And Care NHS Foundation Trust | No | No |
| RP4 | Great Ormond Street Hospital For Children NHS Foundation Trust | No | No |
| RXV | Greater Manchester Mental Health NHS Foundation Trust | No | No |
| RR1 | Heart Of England NHS Foundation Trust | No | No |
| R1A | Herefordshire And Worcestershire Health And Care NHS Trust | No | No |
| RY4 | Hertfordshire Community NHS Trust | No | No |

| | | | |
|---|---|---|---|
| RWR | Hertfordshire Partnership University NHS Foundation Trust | No | No |
| RY9 | Hounslow And Richmond Community Healthcare NHS Trust | No | No |
| RV9 | Humber Teaching NHS Foundation Trust | No | No |
| RXY | Kent And Medway NHS And Social Care Partnership Trust | No | No |
| RYY | Kent Community Health NHS Foundation Trust | No | No |
| RW5 | Lancashire & South Cumbria NHS Foundation Trust | No | No |
| RGD | Leeds And York Partnership NHS Foundation Trust | No | No |
| RY6 | Leeds Community Healthcare NHS Trust | No | No |
| RT5 | Leicestershire Partnership NHS Trust | No | No |
| RY5 | Lincolnshire Community Health Services NHS Trust | No | No |
| RP7 | Lincolnshire Partnership NHS Foundation Trust | No | No |
| RBQ | Liverpool Heart And Chest Hospital NHS Foundation Trust | No | No |
| REP | Liverpool Women'S NHS Foundation Trust | No | No |
| RRU | London Ambulance Service NHS Trust | No | No |
| RW4 | Mersey Care NHS Foundation Trust | No | No |
| RRE | Midlands Partnership University NHS Foundation Trust | No | No |
| RP6 | Moorfields Eye Hospital NHS Foundation Trust | No | No |
| RMY | Norfolk And Suffolk NHS Foundation Trust | No | No |
| RY3 | Norfolk Community Health And Care NHS Trust | No | No |
| RNL | North Cumbria University Hospitals NHS Trust | No | No |
| RX6 | North East Ambulance Service NHS Foundation Trust | No | No |
| RAT | North East London NHS Foundation Trust | No | No |
| G6V2S | North London NHS Foundation Trust | No | No |
| RLY | North Staffordshire Combined Healthcare NHS Trust | No | No |

| | | | |
|---|---|---|---|
| RX7 | North West Ambulance Service NHS Trust | No | No |
| RTV | North West Boroughs Healthcare NHS Foundation Trust | No | No |
| RP1 | Northamptonshire Healthcare NHS Foundation Trust | No | No |
| RHA | Nottinghamshire Healthcare NHS Foundation Trust | No | No |
| RNU | Oxford Health NHS Foundation Trust | No | No |
| RPG | Oxleas NHS Foundation Trust | No | No |
| RT2 | Pennine Care NHS Foundation Trust | No | No |
| R0C | Project Nightingale NHS Trust | No | No |
| RPC | Queen Victoria Hospital NHS Foundation Trust | No | No |
| RXE | Rotherham Doncaster And South Humber NHS Foundation Trust | No | No |
| RAN | Royal National Orthopaedic Hospital NHS Trust | No | No |
| RGM | Royal Papworth Hospital NHS Foundation Trust | No | No |
| TAH | Sheffield Health & Social Care NHS Foundation Trust | No | No |
| R1D | Shropshire Community Health NHS Trust | No | No |
| R1C | Solent NHS Trust | No | No |
| RYE | South Central Ambulance Service NHS Foundation Trust | No | No |
| RYD | South East Coast Ambulance Service NHS Foundation Trust | No | No |
| RV5 | South London And Maudsley NHS Foundation Trust | No | No |
| RYQ | South London Healthcare NHS Trust | No | No |
| RW9 | South Of Tyne And Wearside Mental Health NHS Trust | No | No |
| RQY | South West London And St George'S Mental Health NHS Trust | No | No |
| RXG | South West Yorkshire Partnership NHS Foundation Trust | No | No |
| RYF | South Western Ambulance Service NHS Foundation Trust | No | No |
| RW1 | Southern Health NHS Foundation Trust | No | No |

| | | | |
|---|---|---|---|
| RXX | Surrey And Borders Partnership NHS Foundation Trust | No | No |
| RDR | Sussex Community NHS Foundation Trust | No | No |
| RX2 | Sussex Partnership NHS Foundation Trust | No | No |
| RNK | Tavistock And Portman NHS Foundation Trust | No | No |
| RVX | Tees And North East Yorkshire NHS Trust | No | No |
| RX3 | Tees, Esk And Wear Valleys NHS Foundation Trust | No | No |
| REN | The Clatterbridge Cancer Centre NHS Foundation Trust | No | No |
| RL1 | The Robert Jones And Agnes Hunt Orthopaedic Hospital NHS Foundation Trust | No | No |
| RPY | The Royal Marsden NHS Foundation Trust | No | No |
| RRJ | The Royal Orthopaedic Hospital NHS Foundation Trust | No | No |
| RET | The Walton Centre NHS Foundation Trust | No | No |
| RKL | West London NHS Trust | No | No |
| RYA | West Midlands Ambulance Service University NHS Foundation Trust | No | No |
| RW8 | West Sussex Health And Social Care NHS Trust | No | No |
| RN1 | Winchester And Eastleigh Healthcare NHS Trust | No | No |
| RY7 | Wirral Community Health And Care NHS Foundation Trust | No | No |
| RX8 | Yorkshire Ambulance Service NHS Trust | No | No |

Abbreviations: AA, andexanet alfa; NHS, national health service; ODS, organisation data service

*Trusts with 24 hour consultant-led emergency care activity reported in the last six months.(7)
